# Wastewater sequencing uncovers early, cryptic SARS-CoV-2 variant transmission

**DOI:** 10.1101/2021.12.21.21268143

**Authors:** Smruthi Karthikeyan, Joshua I Levy, Peter De Hoff, Greg Humphrey, Amanda Birmingham, Kristen Jepsen, Sawyer Farmer, Helena M. Tubb, Tommy Valles, Caitlin E Tribelhorn, Rebecca Tsai, Stefan Aigner, Shashank Sathe, Niema Moshiri, Benjamin Henson, Adam M. Mark, Abbas Hakim, Nathan A Baer, Tom Barber, Pedro Belda-Ferre, Marisol Chacón, Willi Cheung, Evelyn S Cresini, Emily R Eisner, Alma L Lastrella, Elijah S Lawrence, Clarisse A Marotz, Toan T Ngo, Tyler Ostrander, Ashley Plascencia, Rodolfo A Salido, Phoebe Seaver, Elizabeth W Smoot, Daniel McDonald, Robert M Neuhard, Angela L Scioscia, Alysson M. Satterlund, Elizabeth H Simmons, Dismas B. Abelman, David Brenner, Judith C. Bruner, Anne Buckley, Michael Ellison, Jeffrey Gattas, Steven L. Gonias, Matt Hale, Faith Hawkins, Lydia Ikeda, Hemlata Jhaveri, Ted Johnson, Vince Kellen, Brendan Kremer, Gary Matthews, Ronald W. McLawhon, Pierre Ouillet, Daniel Park, Allorah Pradenas, Sharon Reed, Lindsay Riggs, Alison Sanders, Bradley Sollenberger, Angela Song, Benjamin White, Terri Winbush, Christine M Aceves, Catelyn Anderson, Karthik Gangavarapu, Emory Hufbauer, Ezra Kurzban, Justin Lee, Nathaniel L Matteson, Edyth Parker, Sarah A Perkins, Karthik S Ramesh, Refugio Robles-Sikisaka, Madison A Schwab, Emily Spencer, Shirlee Wohl, Laura Nicholson, Ian H Mchardy, David P Dimmock, Charlotte A Hobbs, Omid Bakhtar, Aaron Harding, Art Mendoza, Alexandre Bolze, David Becker, Elizabeth T Cirulli, Magnus Isaksson, Kelly M Schiabor Barrett, Nicole L Washington, John D Malone, Ashleigh Murphy Schafer, Nikos Gurfield, Sarah Stous, Rebecca Fielding-Miller, Richard S. Garfein, Tommi Gaines, Cheryl Anderson, Natasha K. Martin, Robert Schooley, Brett Austin, Duncan R. MacCannell, Stephen F Kingsmore, William Lee, Seema Shah, Eric McDonald, Alexander T. Yu, Mark Zeller, Kathleen M Fisch, Christopher Longhurst, Patty Maysent, David Pride, Pradeep K. Khosla, Louise C. Laurent, Gene W Yeo, Kristian G Andersen, Rob Knight

## Abstract

As SARS-CoV-2 continues to spread and evolve, detecting emerging variants early is critical for public health interventions. Inferring lineage prevalence by clinical testing is infeasible at scale, especially in areas with limited resources, participation, or testing/sequencing capacity, which can also introduce biases. SARS-CoV-2 RNA concentration in wastewater successfully tracks regional infection dynamics and provides less biased abundance estimates than clinical testing. Tracking virus genomic sequences in wastewater would improve community prevalence estimates and detect emerging variants. However, two factors limit wastewater-based genomic surveillance: low-quality sequence data and inability to estimate relative lineage abundance in mixed samples. Here, we resolve these critical issues to perform a high-resolution, 295-day wastewater and clinical sequencing effort, in the controlled environment of a large university campus and the broader context of the surrounding county. We develop and deploy improved virus concentration protocols and deconvolution software that fully resolve multiple virus strains from wastewater. We detect emerging variants of concern up to 14 days earlier in wastewater samples, and identify multiple instances of virus spread not captured by clinical genomic surveillance. Our study provides a scalable solution for wastewater genomic surveillance that allows early detection of SARS-CoV-2 variants and identification of cryptic transmission.

## Introduction

SARS-CoV-2 continues to evolve, producing diverse new lineages^1^. Emerging variants of concern (VOCs) and variants of interest (VOIs) demonstrate increased transmissibility, disease severity, and/or immune escape^2^. Timely and accurate quantification of local prevalence of SARS-CoV-2 variants is thus essential for effective public health measures. However, existing strategies for variant detection based on virus genome sequencing of biospecimens obtained from clinical testing (“clinical genomic surveillance”) are expensive, inefficient, and have sampling bias because of systemic healthcare disparities, particularly in poor and underserved communities^3–5^.

In contrast, PCR-based wastewater surveillance of SARS-CoV-2 RNA is not subject to clinical testing biases and can track temporal changes in overall SARS-CoV-2 prevalence in a region ^6–8^, but cannot identify epidemiological transmission links or monitor virus lineage prevalence, which require genome sequence information. Virus genome sequencing from wastewater (“wastewater genomic surveillance”) has the potential to cost-effectively capture community virus spread^9,10^, acting as a surrogate to clinical surveillance in elucidating lineage geospatial distributions and track emerging SARS-CoV-2 variants (including new variants for which targeted assays do not yet exist), and provide genome sequence data needed for transmission network analysis and interpretation^11^.

However, wastewater genomic surveillance is technically challenging^10^. Low viral loads, heavily fragmented RNA, and PCR inhibitors in complex environmental samples lead to poor sequencing coverage^12,13^. Obtaining high quality sequences from samples with low viral load and elevated levels of PCR inhibitors remains an outstanding technical challenge in implementation of wastewater genomic surveillance at scale. Additionally, tools for SARS-CoV-2 lineage classification, such as pangolin^14^ and UShER^15^, were designed for clinical samples containing a single dominant variant, and cannot estimate relative abundances of multiple SARS-CoV-2 lineages in samples with virus mixtures such as wastewater.

Here, we report a high-resolution approach to study community virus transmission using wastewater genomic surveillance, leveraging several technical advances in wastewater virus concentration and nucleic acid sequencing, and a computational tool for resolving multiple SARS-CoV-2 lineages in short-read sequence data from a mixed sample (lineage deconvolution). We obtained near 95% genome coverage even for samples with low viral load, compared with 40% or below from previous studies^11-13^, a key advance that allowed us to build a robust pipeline to monitor virus lineage prevalence in community wastewater.

Because places of communal living, such as university campuses, are considered key sites for virus spread and represent well-controlled and relatively isolated environments, they are ideal for comparing the relative utility of clinical and wastewater genomic surveillance^16^. Accordingly, we conducted a high-resolution, longitudinal wastewater genomic surveillance effort at the University of California San Diego (UCSD) campus, in parallel with clinical genomic surveillance from nasal swabs in the local community, from November 2020 to September 2021: ten months that effectively capture the surges in the region caused by the three main VOCs (as determined by US CDC) in the United States, Epsilon, Alpha and Delta^1^. In more recent San Diego-wide data collected from September 2021 to February 2022, we studied ongoing transmission of the Delta variant and the rapid spread of the Omicron variant and its sublineages.

Our wastewater genomic surveillance approach identified VOCs up to 2 weeks prior to detection through clinical genomic surveillance, even though a large proportion of clinical SARS-CoV-2 samples are sequenced in San Diego relative to other cities in the United States. In addition to providing a detailed history of community virus spread, wastewater genomic surveillance also identified multiple instances of cryptic community transmission not observed through clinical genomic surveillance. Matching wastewater and clinical genome sequences provided epidemiological information identifying specific transmission events. Our results demonstrate the viability of wastewater genomic surveillance at scale, enabling early detection and tracking of virus lineages and guiding clinical genomic surveillance efforts. This work informed public health guidance and interventions on the UCSD campus as well as San Diego county in real time, and our data and analyses were disseminated to both public health officials as well as the general public via custom dashboards (see **Data Availability** for links).

## Results

To directly compare wastewater genomic surveillance to clinical surveillance, we conducted a large-scale SARS-CoV-2 genome sequencing study from wastewater samples collected daily from 131 wastewater samplers covering 360 campus buildings, in many cases reaching single building-level resolution. To identify epidemiological transmission links and monitor lineages in the population, we sequenced all SARS-CoV-2 positive clinical and wastewater samples from campus using a miniaturized tiled-amplicon sequencing approach. During this period of this study, we collected and analyzed 21,383 wastewater samples: 19,944 wastewater samples from the UCSD campus, and, for comparison, 1,475 wastewater samples from the greater San Diego area, including the Point Loma wastewater treatment plant (the primary wastewater treatment plant for the county with a catchment size of 2.3 million people) and 17 public schools spanning four San Diego school districts^17^. We compared sequencing of 600 campus wastewater samples to 759 genomes obtained from campus clinical swabs (46.2% of all positive tests on campus), all processed by the CALM and EXCITE CLIA labs at UCSD. In addition, we compared 31,149 genomes obtained from clinical genomic surveillance of the greater San Diego community to sequencing of 837 wastewater samples collected from San Diego county (including those from the UCSD campus) during the same period.

### High-resolution spatial sampling reveals micro-scale community spread

We implemented a GIS (geographic information system)-enabled building-level wastewater surveillance system to cover 360 buildings on the UCSD campus (**Figure 1A**). During the period of daily wastewater sampling, approximately 10,000 students lived on campus and 25,000 individuals were on campus on a daily basis. We found that wastewater test positivity correlated strongly with the number of clinical positives (**Figure 1B** and **Extended Data Figure 1**), showing that wastewater effectively captures the community infection dynamics based on total viral load. This is also consistent with our past studies that showed SARS-CoV-2 RNA can be detected ∼85% of the time downstream from buildings containing individuals known to be infected^9^.

**Figure 1:**
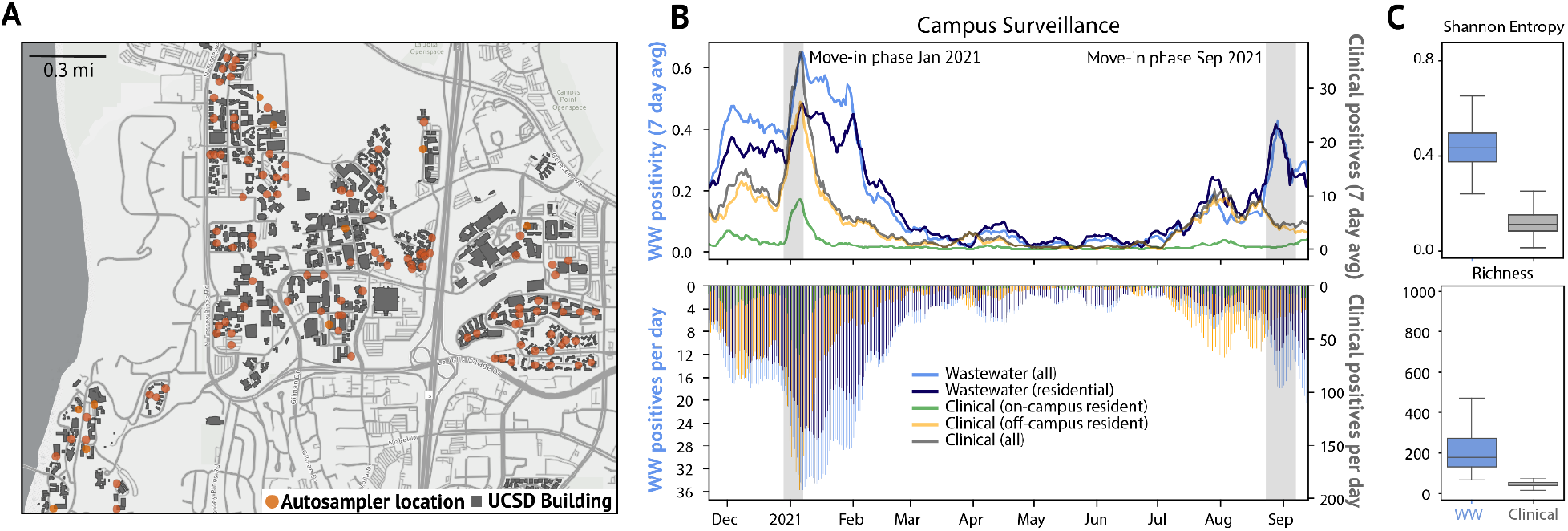
Campus sampling locations and SARS-CoV-2 testing statistics. A. Geospatial distribution of the 131 actively deployed wastewater autosamplers and the corresponding 360 university buildings on the campus sewer network. Building-specific data have been de-identified in accordance with university reporting policies. B. Campus wastewater and diagnostic testing statistics over the 295 day sampling period (WW = wastewater, positivity is the fraction of WW samplers with a positive qPCR signal). C.Virus diversity in wastewater and clinical samples: Boxplots of Shannon entropy (top) and richness (bottom) for each sample type.

Unlike qPCR-based mutant surveillance, genomic surveillance using full-length virus genomes can detect which strains of SARS-CoV-2 are circulating in the population, and can identify potential transmission links between infected individuals^18,19^. While targeted qPCR mutant panels have the ability to detect specific lineages in wastewater, they only target a small set of mutations that must be known beforehand in addition to the development and validation time before implementation. Furthermore, they cannot provide sub-lineage resolution (for instance, BA.1 v. BA.2 sublineage of Omicron) and will fail altogether if a sublineage loses the specific mutation targeted by the qPCR assay. To test the utility of wastewater genomic surveillance for studying virus spread in the community, we obtained near complete virus genomes for wastewater samples with cycle quantification (Cq) values as high as 38 (median genome coverage: 96.49% [75.67% - 100.00%], **Extended Data Figure 2**). However, using two common metrics of virus diversity, Shannon entropy (a measure of the uncertainty associated with randomly sampling an allele) and richness (the number of single nucleotide variant, or SNV, sites)^20^, we found that SARS-CoV-2 genetic diversity is significantly greater in wastewater samples than clinical samples (**Figure 1C**, Mann-Whitney U test, p<0.001 for each, with effect size r=0.99, 0.97 for Shannon Entropy and Richness, respectively). This suggests that multiple virus lineages, likely shed from different infected individuals, are often present in wastewater samples.

### Sample deconvolution robustly recovers the abundance of SARS-CoV-2 lineages in mixed samples

Wastewater systems aggregate stool, urine, and other biological waste products carrying viruses from multiple infected individuals in the community in a single location, allowing for sampling of virus mixtures that are representative of local lineage prevalence. However, existing methods for determining virus lineage from sequencing are intended for non-mixed clinical samples and can only be used to identify a single (dominant) lineage per sample.

To fully capture the virus diversity in community biospecimens, we developed Freyja, a tool to estimate the relative abundance of virus lineages in a mixed sample. Freyja uses a “barcode” library of lineage-defining mutations to represent each SARS-CoV-2 lineage in the global phylogeny^21^(**Figure 2A**). To encode each sample, Freyja stores the SNV frequencies (proportion of reads at a site that contain the SNV) for each of the lineage-defining mutations (**Figure 2B, top**). Since SNV frequencies at positions with greater sequencing depth more accurately estimate the true mutation frequency, Freyja recovers relative lineage abundance by solving a depth-weighted least absolute deviation regression problem, a mixed sample analog of minimizing the edit distance between sequences and a reference (**Figure 2B, bottom**). To ensure results are meaningful, Freyja constrains the solution space such that each lineage abundance value is non-negative, and overall lineage abundance sums to one. Importantly, Freyja performs site-specific weighting to account for non-constant variance in measured SNV frequency across sites, enabling prioritization of information at each site as a function of sequencing depth. Read depths are log-transformed, providing robustness to common attributes of real sequencing data such as heavily skewed read depth across amplicons.

**Figure 2:**
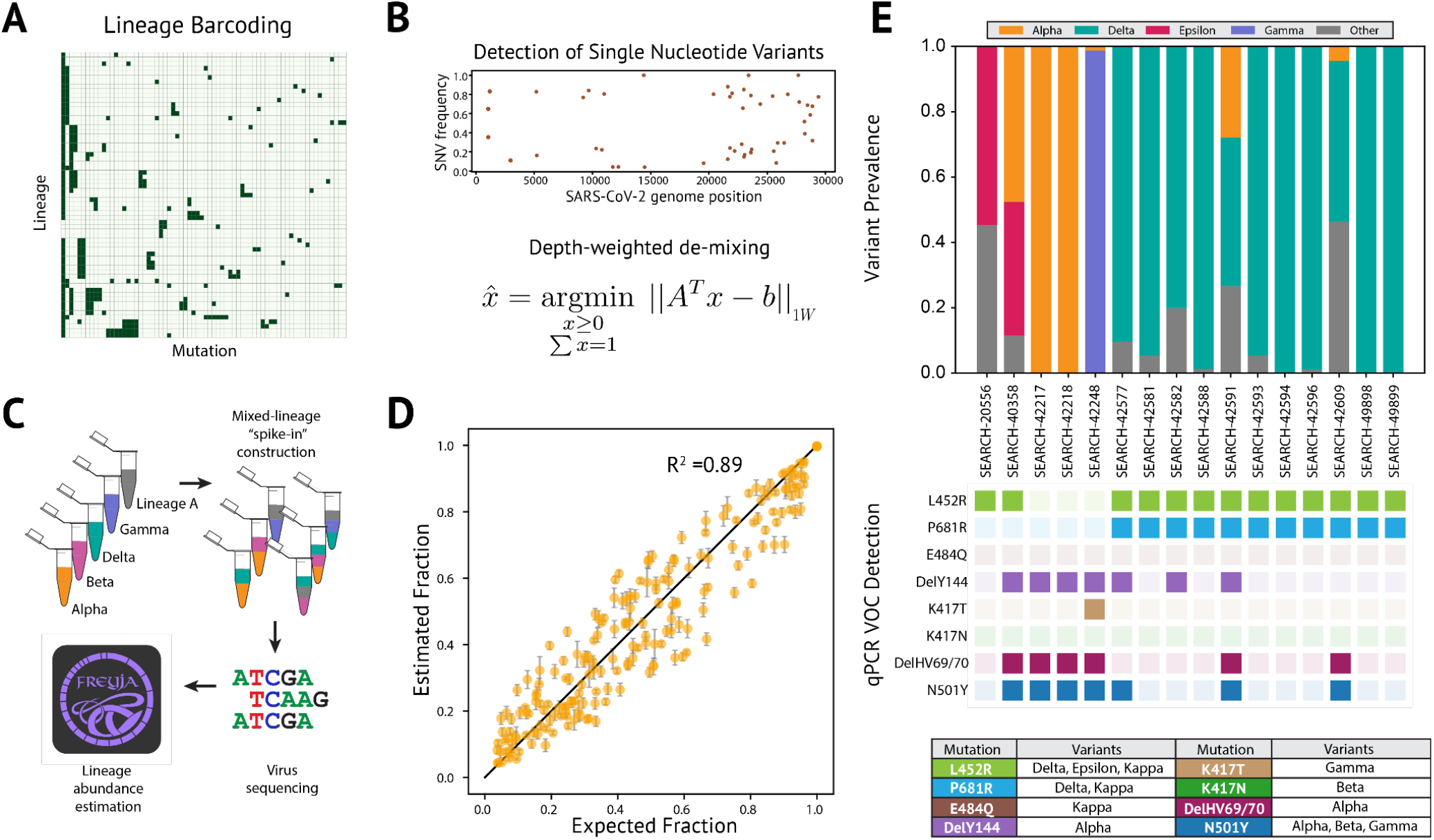
Sample deconvolution robustly recovers relative virus abundance. A. Subset of lineage defining mutation “barcode” matrix. Each row represents one lineage (out of >1000 lineages included in the UShER global phylogenetic tree), and individual nucleotide mutations are represented as columns. B. Single nucleotide variant frequencies obtained from iVar used for recovering relative abundance of each lineage. C. Schematic of the spike-in validation experiment. D. Depth-weighted de-mixing estimates of the virus abundance versus expected/known abundance. Details on lineage specific predictions are provided in **Extended Data Figure 3**. E. Comparison of wastewater sample deconvolution with VOC qPCR panel, with lookup table (bottom) showing amino acid mutations corresponding to each variant.

To validate Freyja, we sequenced “spike-in” synthetic mixtures from five key SARS-CoV-2 lineages (Lineage A, Beta, Delta, Epsilon, and Gamma) at proportions ranging from 5% to 100% in each sample, with between 1 and 5 different lineages per mixture (**Figure 2C**, and see **Table 1**). We found that Freyja robustly recovered the expected lineage abundances for all mixtures, even for lineages at 5% abundance (**Figure 2D**, and see **Extended Data Figure 3** for lineage specific predictions).To further validate Freyja, we used wastewater samples from the UCSD isolation dorms as well as Point Loma wastewater treatment plant, collection sites likely to contain mixed-lineage samples, to compare Freyja-detected lineages with qPCR testing for 8 mutations associated with different variants of concern (N501Y, DelHV69/70, DelY144, K417N, K417T, E484Q, P681R and L452R, **Figure 2E**). We found that Freyja consistently identified the same lineages as qPCR testing, but, as expected, also identified additional lineages with SNVs not included in our qPCR panel that were known to be circulating in San Diego at the time of collection. Combined, these results show that Freyja robustly estimates viral lineage abundance from samples containing a mixture of lineages, including synthetic virus mixtures and field wastewater collections.

To compare Freyja with other wastewater analysis pipelines, we tested the performance of other wastewater deconvolution methods including the method from Baaijens et al.^12^, cojac^22^, and LCS ^23^ using the spike-in mixtures (**Extended Data Figure 4)**. We found that Freyja greatly outperforms other methods in terms of accuracy, false positive rate, and computational efficiency. The method from Baaijens et al. required greater than ten times more computation time per sample relative to Freyja (∼13.2 minutes vs ∼1.1 minutes per sample, respectively). Although cojac was fast, the small amplicon length used for the spike-in mixtures caused cojac to fail to identify most of the variants entirely, while LCS failed to return estimates within two days.

### Detection of early and cryptic community transmission in wastewater

SARS-CoV-2 RNA concentrations in wastewater have been shown to be an early indicator of rising COVID-19 community incidence^9,24^ (and see **Extended Data Figure 5A**), but whether wastewater can be used to detect emerging variants, including VOCs and VOIs, prior to their observation in clinical surveillance is unknown. To test if wastewater can enable early detection of emerging lineages, we applied Freyja to our wastewater sequencing data and compared the collection date of VOC positive samples from wastewater with the collection dates of samples from clinical genomic surveillance (**Figure 3A**). With only 2.6% as many sequenced wastewater samples as sequenced clinical samples, we detected the Alpha and Delta VOC lineages in wastewater genomic surveillance up to 14 days prior to their first detection in genomic clinical surveillance (Epsilon was circulating at the start of wastewater collection, and thus could not be detected early). To further quantify our uncertainty in prevalence estimates, we used a fast bootstrapping approach (**Extended Data Figure 6)** and found that the resampled distributions did not include zero abundance. Since emerging VOC lineages may evade immune responses or lessen the effectiveness of public health interventions^18^, this early detection provides additional time to make necessary adjustments to existing countermeasures.

**Figure 3:**
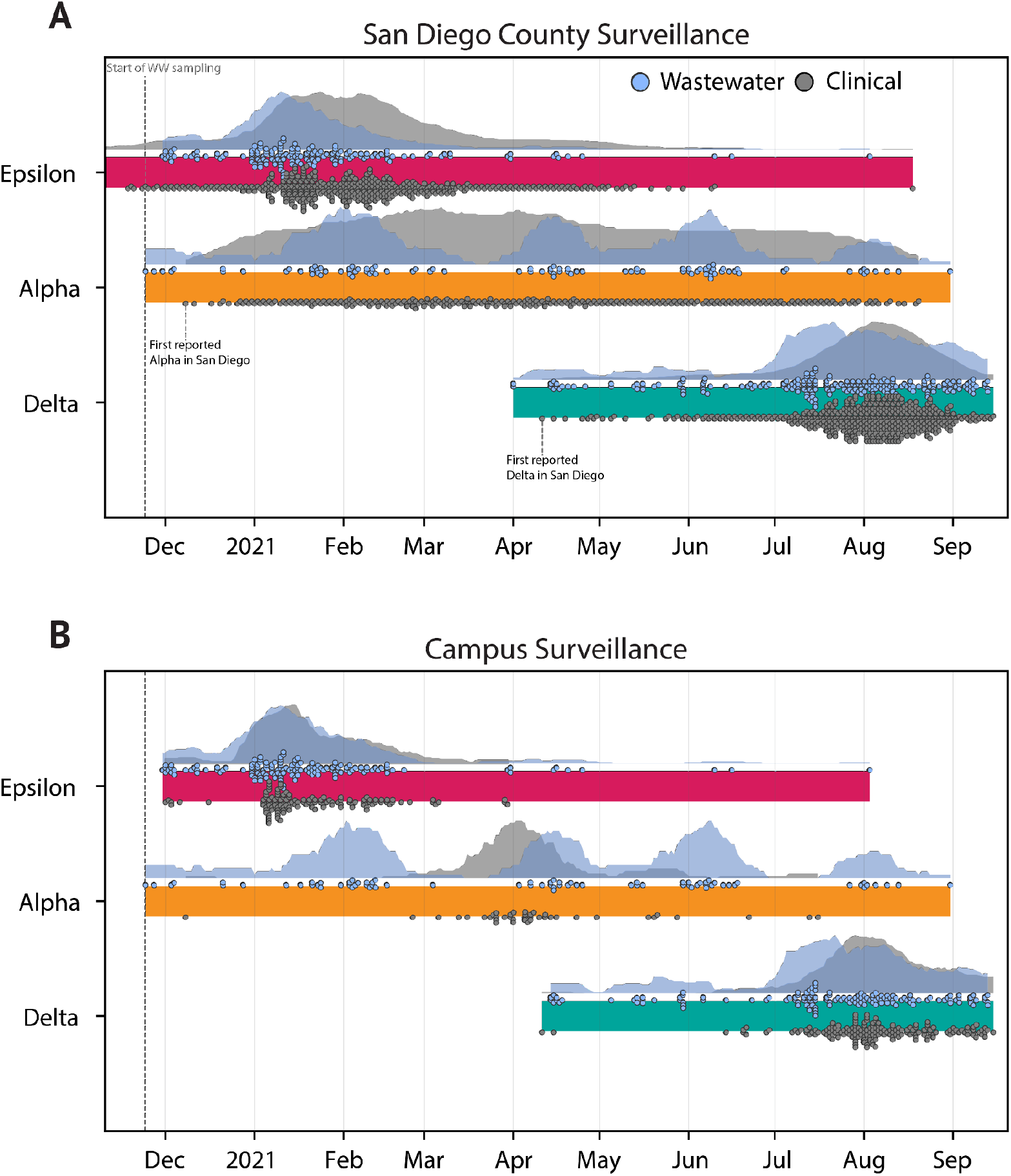
Freyja recovers early and cryptic transmission of SARS-CoV-2 variants of concern. A. Timeline and normalized epidemiological curves for VOC detection in both wastewater and clinical sequences from San Diego County for the 3 major VOCs in circulation during the sampling period. Both Alpha and Delta are detected first in wastewater before clinical samples. Markers for clinical detections correspond to the ceiling of the daily detection count divided by 30 (e.g. 1-30 samples= one marker, 31-60 = two markers), while wastewater markers correspond to a single detection. B. Timeline and epidemiological curves for VOC detection in the campus samples. Markers correspond to a single detection event for both clinical and wastewater surveillance. All wastewater detections correspond to an estimated VOC prevalence of at least 10%.

To test if wastewater genomic surveillance can identify changes in the abundance of circulating lineages, we compared VOC detection rates in clinical and wastewater sequencing over time. We found that both wastewater and clinical genomic surveillance tracked changes in lineage abundance, but increases in lineage detection frequency were generally observed first in wastewater surveillance. For example, for the Epsilon variant, which was first detected in San Diego in September of 2020, we observed increases in detection frequency in wastewater approximately 5 days prior to the corresponding increase in clinical genomic surveillance data (**Figure 3A**, see **Methods**). We noticed varying periods of ongoing lineage detection across VOCs relative to clinical surveillance, possibly due to different virus shedding characteristics across lineages ^25^. For Epsilon specifically, elevated sampling density on the UCSD campus relative to elsewhere in the county early on in the experiment may have biased San Diego wide detection trends towards campus trends, particularly during the end of the wave. We also observed clear signatures of times with elevated travel, as seen in the pulsing of Alpha detections in wastewater around the end of holidays and school breaks. During these periods as well as other times of mass student arrival, students were mandated to test immediately upon arrival before they moved into their respective on-campus housing. In late March of 2021 following the university break, mandated clinical testing identified spread of the Alpha variant exclusively in off-campus residents (see **Figure 1B)**, suggesting that campus mitigation protocols kept the Alpha outbreak from spreading on campus during this period.

To study the effectiveness of wastewater genomic surveillance at a smaller community scale, we restricted our analysis to samples from the UCSD campus. We found that wastewater genomic surveillance consistently identified the three major VOCs (Epsilon, Alpha, and Delta) throughout their period of occurrence, despite detection gaps of one month or longer in clinical surveillance that included regular asymptomatic testing, longer than the expected signal due to extended virus shedding ^26–28^ (**Figure 3B**). During these gaps, positive samples were collected from multiple distinct locations, with most locations not repeated, suggesting that this continued detection in wastewater was not simply due to extended shedding. From mid-December to late-March, the Alpha variant was detected more than once per week on average in wastewater but was not detected by clinical surveillance. Similarly, wastewater surveillance detected continued Delta transmission from mid-April to mid-June, but no cases were identified by clinical surveillance. This explains in part the long tails of wastewater positivity on campus relative to clinical surveillance on campus (**Figure 1B)**, in which we control for extended shedding by excluding samples from campus isolation dorms (see **Methods** for details). The high wastewater positivity level in February-March 2020 extends beyond the expected duration of extended shedding, indicating that cryptic transmission likely played a significant role in campus virus spread during this period.

To study the effectiveness of wastewater surveillance in detecting and tracking other emerging variants, we aggregated all wastewater sequencing data to estimate the temporal profile of community lineage prevalence. We found that estimates of lineage abundance using wastewater enable early identification of other VOCs/VOIs, even for lineages that are rarely observed in clinical surveillance (**Figure 4**). For example, we detected the Mu (B.1.621) variant via wastewater genomic surveillance on July 27th, nearly four weeks prior to its first detection through clinical genomic surveillance on campus, on August 23rd (**Figure 4A,C**). However, despite persistent Mu detection in campus wastewater throughout July and early August, we did not detect the Mu variant in clinical or wastewater genomic surveillance on campus in September, suggesting that local community transmission did not continue.

**Figure 4:**
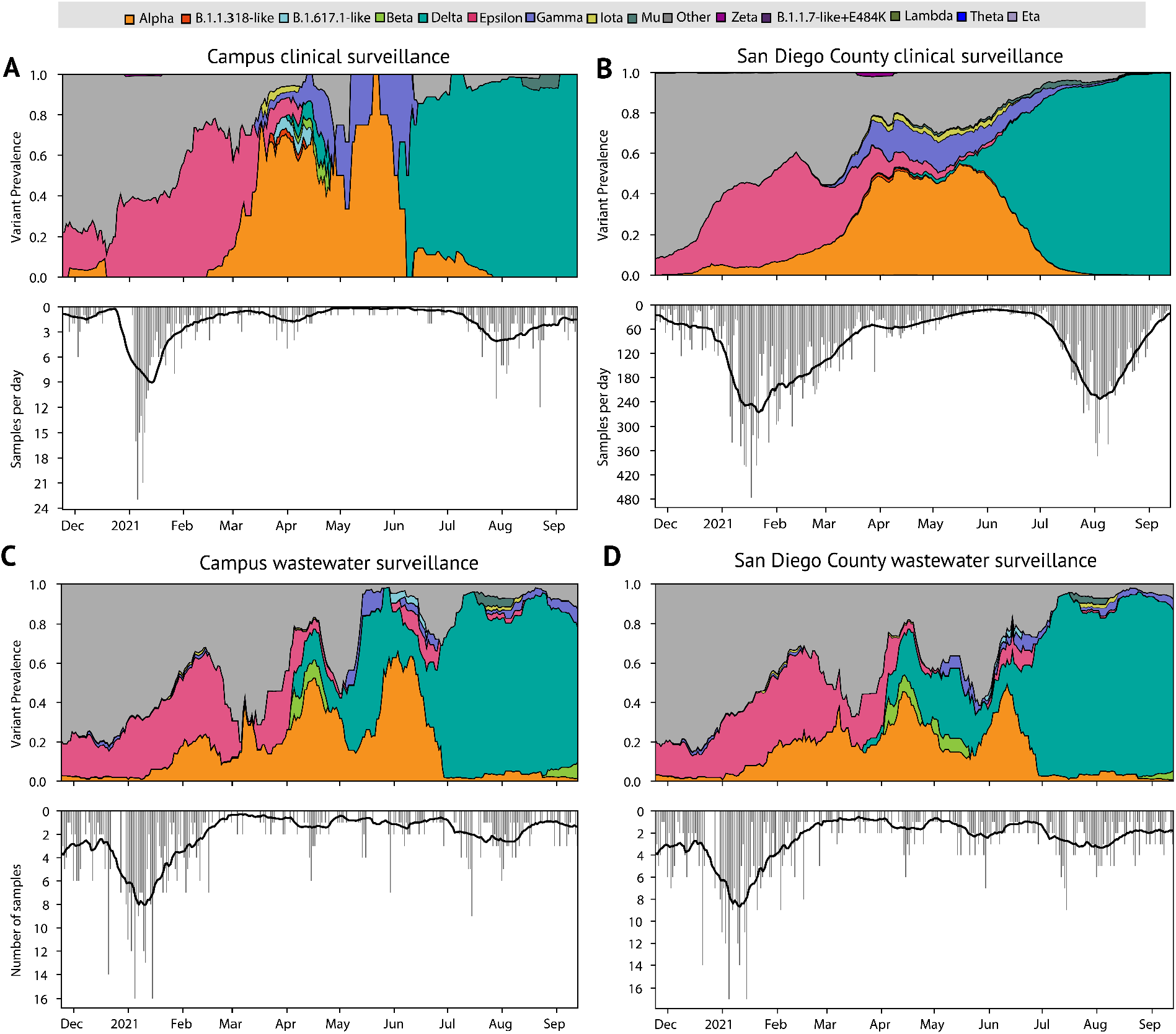
Deconvolution recovers a fine-grained estimate of virus population dynamics. A. Prevalence of SARS-CoV-2 variants in UCSD clinical surveillance, and B. Variant prevalence in all clinical samples collected in San Diego County. C,D. Variant prevalence in wastewater at UCSD as well as the greater San Diego County (includes wastewater samples collected from Point Loma wastewater treatment plant as well as public schools in the San Diego districts). Further analysis of Point Loma wastewater samples is shown in **Extended Data Figure 5**. All curves show rolling average, window ±10 days. “Other” contains all lineages not designated as VOCs. Bottom panels show number of sequenced samples per day.

To test if Freyja continues to provide representative estimates of lineage prevalence for mixtures containing closely related lineages, we analyzed the rise of the Delta variant (B.1.617.2) and its sublineages (AY.*) in San Diego, from June-September 2021 (**Extended Data Figure 5B,C**). At both the UCSD campus and the Point Loma wastewater treatment plant, we identified the rapid emergence of B.1.617.2 and its sublineages (AY.*), along with low but persistent levels of the P.1 (Gamma) variant. The relative abundances of each of the variants were within 2-fold of prevalence estimates observed in clinical nasal swab data, suggesting that Freyja effectively identifies prevalence even for closely related lineages, both at the university and county-scale.

In more recent data from Point Loma wastewater treatment plant, we identified the Omicron variant (B.1.1.529 and descendants) at an abundance of near 1.7 % on November 27th, more than 10 days prior to the first clinical detection in San Diego on December 8th (**Figure 5A-B**). To confirm these findings, we applied our VOC qPCR panel to the same samples and consistently detected two mutations associated with the Omicron variant (DelHV69/70 and N501Y) in samples detected after November 27th, while neither was detected in samples from earlier in November (**Extended Data Table 3**, P681R was included to confirm the presence of Delta).

**Figure 5:**
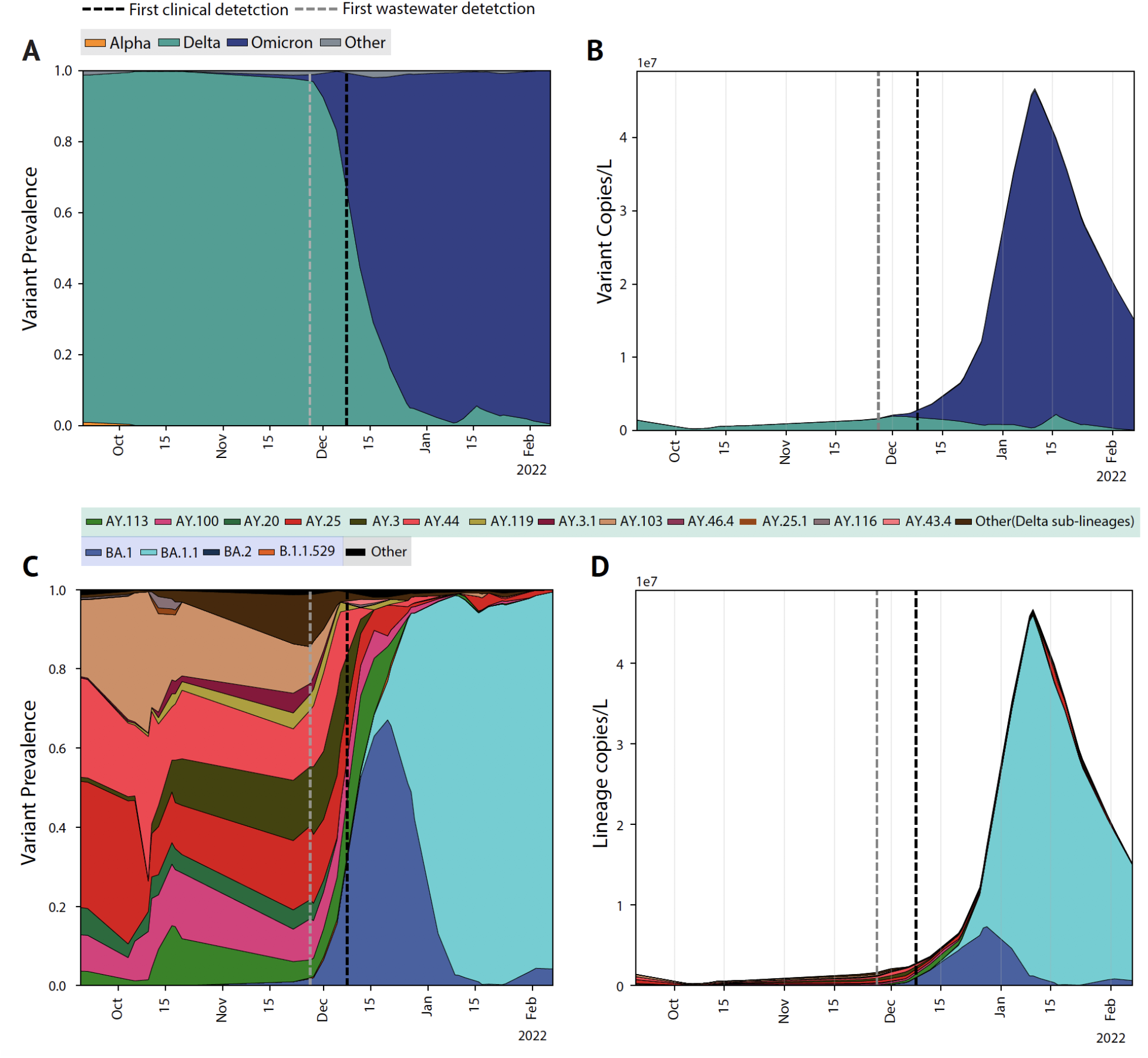
Community wastewater enables early Omicron detection and reveals lineage dynamics. A. Prevalence of SARS-CoV-2 VOCs in wastewater collected from the Point Loma wastewater treatment plant from late September 2021 to early February 2022. B. Estimated VOC concentrations, prevalence estimates scaled by normalized viral load in wastewater. C,D. Lineage-specific estimates of prevalence and concentration. All curves show an adaptive rolling average calculated using a local linear approximation (Savitzky-Golay filter) of virus copies/L, with window size ± 1 sampling date.

To visualize the dynamics of competition between the Delta and Omicron variants, we analyzed wastewater collected at Point Loma from late September through early February. We found that upon introduction to the community, Omicron rapidly rose to dominance and reached roughly 95% prevalence by December 26th. During the same period, the estimates for 95% Omicron abundance in clinical samples tracked via S-gene target failures (SGTFs) was January 7th, further suggesting wastewater genomic surveillance is a leading indicator of lineage dynamics for emerging variants (**Figure 5A, Extended Data Figure 7**). To understand the magnitude of lineage abundance, we scaled each sample by the measured virus RNA concentration of the sample (**Figure 5B**). We observed that the absolute amount of circulating Delta variant remained largely constant upon the introduction of Omicron, even as it appeared to decrease to a small fraction of all viruses in the community.

To study the contribution of individual virus lineages to virus RNA concentration, we further analyzed the growth dynamics of Delta and Omicron sub-lineages (**Figure 5C-D**). We found that the many Delta lineages circulating in October and November were rapidly displaced by the BA.1 Omicron lineage, which was soon after displaced by the BA.1.1 lineage, suggesting a significant growth advantage over BA.1 and B.1.1.529. We did not observe significant levels of any other Omicron sublineages.

### Wastewater identifies both known and unknown history of campus infections

Phylogenetic analysis of virus genomes can be used to identify fine-scale spatial and temporal transmission networks, but it is unknown if wastewater can be used to further refine possible sites of transmission, elucidate transmission networks (“who-infected-whom”), or identify specific infected individuals^19^. To investigate the scale, structure, and timing of SARS-CoV-2 spread on campus, we reconstructed a maximum likelihood phylogenetic tree for each of the major VOCs using all high-quality consensus genomes (see **Methods** for details) obtained from the UCSD campus, as well as reference sequences for each lineage obtained elsewhere in the United States (**Figure 6A-C**). In each tree, we identified many independent introductions, some of which led to extended transmission on campus. The resulting virus diversity among the VOCs present on campus enables ruling out of most transmission links and suggests campus virus spread consisted of many separate, small outbreaks.

**Figure 6:**
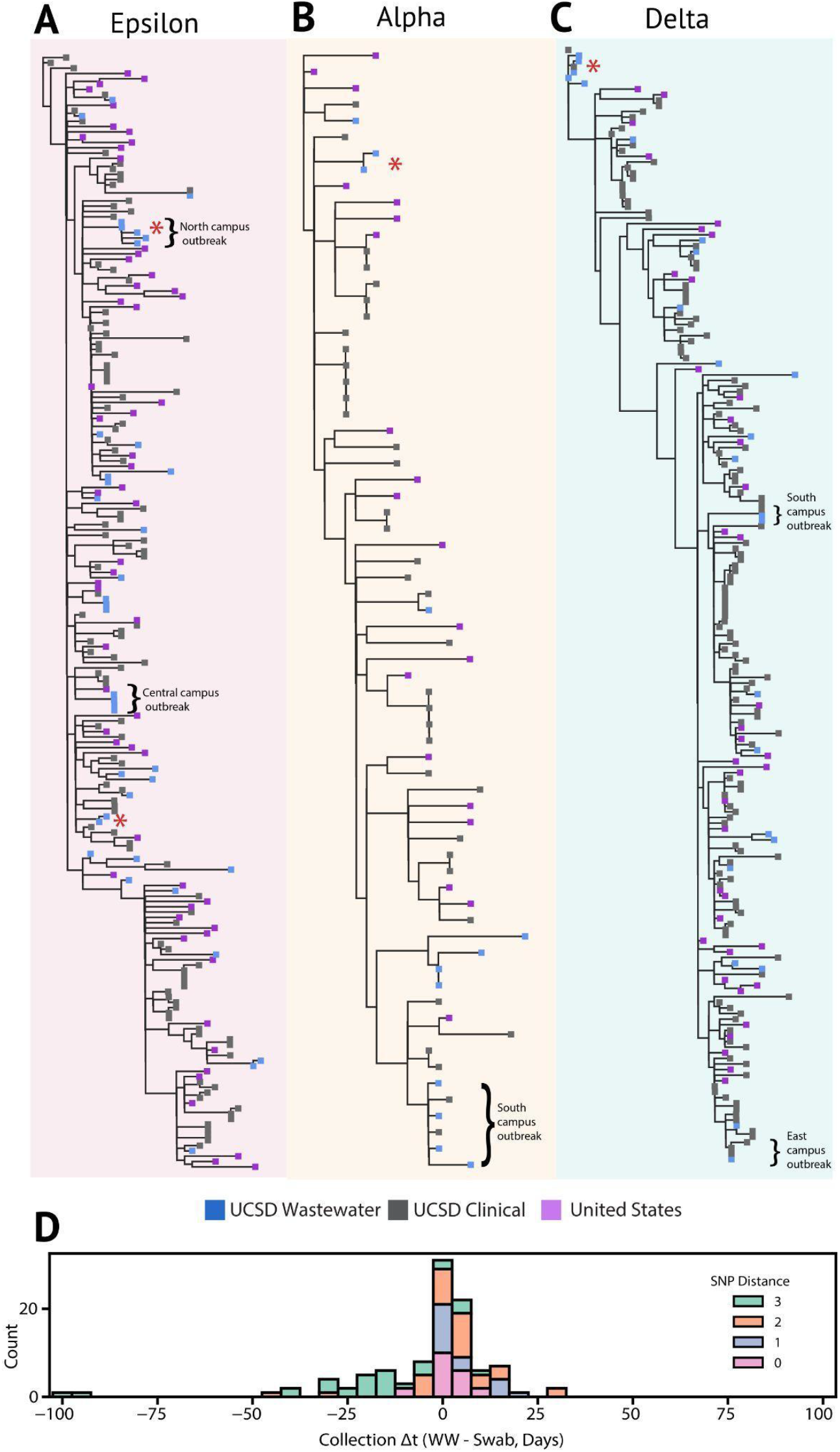
Wastewater identifies clinically known and unknown virus transmission. A-C. Maximum likelihood phylogenetic trees for each of the dominant variants of concern using high quality samples obtained at UCSD, as well as a representative set of sequences from the entire United States. Wastewater sequences from the same sampler that differ by 1 or fewer SNPs are denoted with a red asterisk. For all sequences, consensus bases were called at sites with >50% nucleotide frequency. Location information is provided for select outbreaks. D. Pairwise comparison of collection date for matching and near-matching wastewater and nasal swab samples obtained at UCSD. Positive values indicate earlier collection in nasal swabs, and negative values indicate earlier detection in wastewater.

To analyze the spatial structure of virus spread, we identified collection sites for wastewater sequences connected to transmission chains on campus, with building-specific resolution (**Figure 6 A-C**). We observed multiple small, linked outbreaks clustered in nearby buildings. Campus isolation protocol required students in congregate living to relocate to an isolation room and linkages in the wastewater samples from buildings used for isolation reflected this co-location. We also found multiple instances of successive exactly matching sequences from wastewater collected from a single building, possibly due to continued viral shedding from the same infected individuals from extended shedding in stool^26–28^ or a transmission chain in the building leading to multiple infections by genetically identical viruses.

To study the temporal delay between clinical and wastewater lineage detection, we compared collection times of sequences from campus wastewater that match sequences from campus clinical surveillance (including non-VOC lineages). We found 20 exact sequence matches and 103 near-matches (SNP distance of 3 or less) but did not observe any overall bias towards earlier or later detection in wastewater (**Figure 6D**), suggesting that on average, wastewater and clinical genomic surveillance identify a similar timing of individual detection events. However, despite current technical difficulties with isolating haplotypes from diverse virus mixtures, more than half of the clinical-wastewater sequence pairs demonstrate earlier detection in wastewater or are from the same date. Importantly, since detection is often delayed or missed by clinical surveillance, detections occur first in wastewater (despite a loss of sequences due to limited haplotype recovery), further suggesting that wastewater genomic surveillance can reveal the presence of specific genome sequences prior to clinical surveillance.

## Discussion

We show that improved virus concentration from wastewater, coupled with a method for resolving multiple lineages from mixed samples, captures community virus lineage prevalence and enables early detection of emerging variants, often before observation in clinical surveillance. By sequencing both clinical and wastewater samples from the UCSD campus, we detect VOCs persistently in wastewater even when their appearance in clinical samples is intermittent. However, we also found occasions when rarer lineages, like B.1.1.318, were detected in clinical samples but not in wastewater. This is not unexpected on campus since many students living off-campus did not contribute to campus wastewater but were still clinically tested as part of testing mandates and policies. In the larger San Diego community context, this suggests that we may not be able to identify lineages circulating at low prevalence (< 1%) using a single wastewater collection site. In addition, we note that clinical sequences identified from the community may not be observable in the contributing catchment, as precise geolocation of all clinical samples was not possible. On the other hand, we also observed rare lineages in wastewater not seen in clinical samples from campus or the community. Since campus testing mandates are unable to capture all cases (e.g. fully vaccinated individuals were not required to test and not all community samples were sequenced), rare lineages can be missed.

The considerable benefits of wastewater surveillance may stem from biases in clinical testing, including population testing availability and compliance, university quarantine policies, and asymptomatic transmission, which may distort estimates of virus lineage prevalence from clinical samples. Wastewater offers less biased and more consistent viral lineage prevalence estimates, especially in areas with limited access and/or higher testing hesitancy rates, where limited clinical surveillance can delay detection of emerging variants. Since it requires considerably fewer samples, it is also more cost-effective than clinical testing, and could serve as a long-term passive surveillance tool. This is particularly important for developing public health interventions in low-resource and underserved communities, where widespread clinical genomic surveillance for SARS-CoV-2 remains limited.

Wastewater is an information-dense resource for estimating the prevalence of specific viral lineages, providing a community wide-snapshot not only of overall infection dynamics but of the rise and fall of specific VOCs. Our method, Freyja, deconvolutes these information-rich mixtures of virus lineages. For a large catchment area, such as San Diego’s Point Loma wastewater treatment plant, which covers over 2 million residents, even limited sampling may accurately estimate lineage prevalence in the population and provide an early warning indicator of the rise of new VOCs (as evidenced by the detection of Omicron at just over 1% abundance 11 days ahead of the first local clinical observation). In addition, wastewater genomic surveillance with building-level resolution provides a detailed description of the structure and dynamics of community virus transmission, and can identify transmission links. It can be used to better direct public health interventions, and can do so in real-time when combined with fast-turnaround sequencing technologies. This high-resolution approach is of particular utility in community gathering and transit sites, such as schools and airports, as well as sites with highly vulnerable individuals, such as nursing homes and hospitals, where spatially resolved monitoring for directing public health interventions is of great importance.

As SARS-CoV-2 continues to evolve, the risk of new VOCs remains high and there is a growing need to identify these viruses ahead of their proliferation in the community. Accordingly, development of technologies that are cost-effective, reduce biases, and provide leading rather than trailing indicators of infection are essential to removing “blind spots” in our understanding of local virus dynamics. Although technical issues have made wastewater sequencing difficult to perform at scale, our key advances in virus concentration and sample deconvolution provide evidence that this approach is now viable. Continued improvements to sequencing turnaround speeds, lineage barcoding, and haplotype recovery from mixed samples will further accelerate efforts to achieve earlier identification of emerging variants and improve the precision and effectiveness of interventions.

## Methods

### Wastewater sampling

#### High-resolution spatial sampling at the campus level

131 wastewater autosamplers collecting 24h time-weighted composites were deployed across manholes or sewer cleanouts of 360 campus buildings. GIS (geographic information systems) informed analyses as well as agent-based network modeling of SARS-CoV-2 transmission on the UCSD campus enabled identification of most optimal locations for wastewater sampling. During the pilot phase (November 23-Dec 29^th^ 2020), 68 samplers were prioritized to cover 239 residential buildings identified as the highest risk areas for large outbreaks on campus as a part of an observational study of wastewater monitoring in high-density buildings ^29^. This was based on preliminary dynamic modeling which showed the largest potential outbreaks to occur within the largest residential buildings ^9^. In addition to the observational study of wastewater monitoring in these high-density buildings, a cluster randomized study was also performed concurrently. This included a randomized modified version of a stepped wedge crossover design, in which there was random assignment of manholes for wastewater sampling. Clusters of manholes associated with residential buildings were randomized to receive wastewater monitors at one of two-time steps to evaluate the impact of wastewater monitoring on outbreak size in the associated buildings. During the same time period, all students in these residences were mandated to undergo weekly diagnostic testing which was used to validate the utility of building-level wastewater monitoring. Furthermore, on-campus residences were initially focused due to the relatively static nature of the population which enabled a more robust cross-validation of the sensitivity and efficacy of the wastewater surveillance. The coverage of wastewater surveillance was then increased to cover the rest of the campus buildings (including non-residential buildings on campus) from January 2021. Four of the deployed wastewater samplers covered the designated isolation and quarantine buildings on campus.

Wastewater composites were collected from the 131 samplers every day for the on-campus residence buildings and Monday through Friday for the nonresidential campus buildings. 19,944 wastewater samples were collected and analyzed for the presence of SARS-CoV-2 RNA via RT-qPCR between November 23^rd^ 2020 and September 20^th^ 2021. During this time, 9700 students lived in campus residences and 25,000 worked on campus on a daily basis. Between October 2020 to January 1st 2021, all on-campus residents were mandated to test on a bi-weekly (once every 2 weeks) basis and on a weekly basis from January 2nd 2021 (start of the Winter term). However, fully vaccinated individuals were not mandated to test on a regular basis. Campus protocols required SARS-CoV-2 positive students living in congregate housing to relocate to designated isolation housing. Accordingly, our analysis of wastewater positivity (**Figure 1B)** did not include isolation housing samplers, in order to control - as best as possible, a small number of students in non-congregate housing spaces were allowed to isolate “in-place”, for example - for possible repeat detection due to extended shedding from infected individuals. Automated, localized wastewater-triggered notifications were sent to the residents/employees of buildings associated with a positive wastewater signal which further led to a surge in testing uptake rates by 2 to 40-fold in the associated buildings.

#### Wastewater sampling at the county level

24h flow-weighted composites were collected thrice a week from the main pump station for the Point Loma wastewater treatment plant, the primary treatment plant serving the greater San Diego county with a catchment size of approximately 2.3 million. 132 wastewater samples were collected between February 24th 2021 to February 7th, 2022.

### Wastewater sample processing and viral genome sequencing

#### Sample processing

SARS-CoV-2 RNA was concentrated from 10ml of raw sewage and processed as described elsewhere^7^. In brief, the viral RNA was concentrated using an automated affinity capture magnetic hydrogel particle (Ceres Nanosciences Inc., USA) based concentration method after which the nucleic acid was extracted and sample eluted in 50uL of elution buffer. The extracted RNA was then screened for SARS-CoV-2 RNA via real-time RT-qPCR for 3 gene targets (N1, N2 and E-gene). PMMoV (pepper mild mottle virus) was also screened to adjust for changes in load. Positive wastewater samples were sequenced within 1-2 weeks of collection, comparable to the delay for clinical samples. To cross-validate the ability of the deconvolution tool in reliably resolving mixtures of strains in wastewater, the wastewater samples from the county as well as the ones from the isolation dorms on campus (where multiple infected individuals were isolating) were also run through a PCR panel targeting 8 mutations associated with the strains designated as VOCs. The mutations screened for in wastewater using RT-qPCR included N501Y, DelHV69/70, DelY144, K417N, K417T, E484Q, P681R and L452R (Promega Corp. Cat# CS3174B02).

#### Miniaturized wastewater SARS-CoV-2 amplicon sequencing

The Swift Normalase® Amplicon Panels (SNAP) kit (PN: SN-5×296 (core) COVG1V2-96 (amplicon primers), Integrated DNA Technologies, Coralville, IA) was used on RNA from wastewater samples that were positive for SARS-CoV-2 RNA to prepare the multiplex NGS amplicon libraries and indexed using the SN91384 series of dual indexing oligos, yielding up to 1536 index pairs per pool. A miniaturized version of the protocol was used with the following modifications: the Superscript IV VILO (Thermo Fisher, Carlsbad, CA) cDNA synthesis reaction was scaled down to ∼1/12 the normal reaction volume with 0.333uL of enzyme mix and 1.333uL of RNA being used. The multiplex amplicon amplification and Ampure XP bead purification steps were scaled down ∼1/6 the normal reaction volume. The Index adapter PCR reaction and Ampure XP bead purification steps were scaled down to ∼2/13 the normal reaction volume. The final library resuspension volume was 29uL. 1uL of each library was pooled for an initial shallow NGS run on a MiSeq (Illumina, San Diego, CA) using a Nano flow cell. This equal volume pool was used to estimate the differential volumes required for similar read depths across samples using a NovaSeq SP or S4 flow cell (Illumina, San Diego, CA). Between 5uL and 0.2uL of library material, depending on the data provided from the MiSeq Nano run, was pipetted into a single pool for the NovaSeq run. Transfer volumes were capped at 5uL to reduce pipetting time and because these types of “high volume” samples typically contained a higher proportion of likely adapter dimers that inhibit flow cell performance for all samples. A Dragonfly Discovery (SPT Labtech, UK) was used to dispense reaction master mixes or water depending on the step. A BlueWasher (BlueCatBio, MA) was used for high throughput centrifugal 384-well plate washing during the AmpureXP bead reaction cleanup steps. An IKA MS3 Control linear plate mixer (IKA Works Inc, Wilmington, NC) set to 2600 RPM for 5’ was used to resuspend the AmpureXP beads during the rehydration steps. A Mosquito Genomics HV 16 channel robotic liquid handler (SPT Labtech, UK) was used to dispense the RNA, the reaction master mixes, and prepare the equal volume pools for the initial MiSeq Nano (Illumina, San Diego, CA) balancing runs. A Mosquito X1 single channel “hit picker” robotic liquid handler (SPT Labtech, UK) was used for the final library balancing for the NovaSeq (Illumina, San Diego, CA) NGS lanes.

Sequencing data were analyzed using the C-VIEW (COVID-19 VIral Epidemiology Workflow) platform for initial QC and SARS-CoV-2 lineage assignment and phylogenetics. In brief, sequencing reads are aligned with minimap2^30^, and primer sequences trimming and quality filtering is applied using the iVar trim method^20^. Sequencing depth and single nucleotide variant (SNV) calls are obtained using samtools mpileup^31^ and the iVar variants method^20^.

Controls were included at all stages of sample processing (viral concentration, extraction, qPCR and sequencing) to assess potential inhibition and cross-contamination. Most of the sample processing steps were performed by liquid handling robots for consistency and to minimize human error. Replicates were included for all wastewater samples. If any of the controls failed or indicated cross-contamination, the entire batch was rerun. The clinical samples and wastewater samples were processed separately for sequencing due to significant differences in viral load between the two sample types.

### Virus diversity

As reported previously^20^, virus SNVs were used to characterize the populations derived from wastewater and clinical samples. Richness was defined as the total number of SNV sites, and mean Shannon entropy *H*(*p*) was defined as

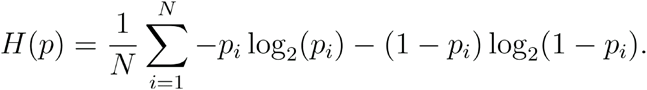

where *p*_*i*_ is the SNV frequency of at the i-th site, of *N* total sites. For statistical testing, a Mann-Whitney U test was performed using all wastewater samples that were not sampled from the same source within a 10 day period in order to ensure independence across samples, as well as all clinical samples. Effect size was calculated using the rank-biserial correlation, *r* = 2*U* / (*n*_*WW*_*n*_*CS*_) − 1where *U* is the Mann-Whitney test statistic and *n*_*WW*_ and *n*_*CS*_ are the numbers of wastewater and clinical samples, respectively.

### Wastewater sample deconvolution

To infer relative abundance within a wastewater sample, we used a “barcode” matrix containing the lineage defining mutations for each known virus lineage,

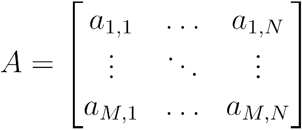

where *a*_*i,j*_ denotes the i-th lineage, at mutation j. Lineage defining mutations were obtained from the UShER global phylogenetic tree using the matUtils package^15^. Similarly, we let *b* and *d* encode the frequency of each mutation and the corresponding sequencing depth (using the log-transform *d*_*i*_ = *log*_2_(depth_*i*_ + 1) to adjust for large differences in depth across amplicons, which we use to control for heteroskedasticity and down-weight the importance of sites with little or no sequencing depth),

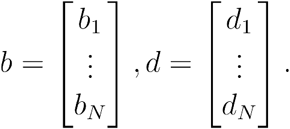

We were then able to write this as a constrained (weighted) least absolute deviations problem

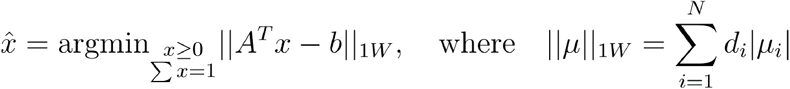

which yields the “demixing” vector 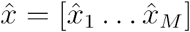 that specifies the relative abundances of each of the known haplotypes. Analysis was only performed on samples with greater than 70% coverage, with the exception of March samples from UCSD for which all samples with greater than 50% coverage were used. Constrained minimization was performed in Python using the cvxpy convex optimization package^32,33^. Mapping of lineages to variant WHO lineages (VOCs, VUMs, etc.) was performed using curated lineage data from outbreak.info^1^. We note that the Epsilon variant received different maximum escalation levels at CDC and WHO, which assigned VOC and VOI status, respectively. Since the Epsilon variant was widespread in California and much of the United States, we use the more “local” CDC designation.

### Fast-bootstrapping method

Bootstrapping was performed at the nucleotide level by resampling each site based on a multinomial distribution of read depth across all sites, where the event probabilities are determined by the fraction of the total sample reads found at each site, followed by a secondary resampling at each site according to a multinomial distribution (i.e. binomial when there was only one SNV at a site), where event probabilities were determined by the frequencies of each base at the site, and the number of trials is given by the sequencing depth. 1000 resamplings and demixings were performed for all samples.

### Spike-in mixture experiment

RNA was isolated from supernatants of a mammalian cell culture infected with one of five strains of SARS-CoV-2. (A, B.1.1.7, B.1.351, P.1, or B.1.617.2).

#### RNA concentration standardization

Virus concentration was quantified by the UCSD EXCITE COVID testing laboratory using the Thermo COVID-19 Test kit (PN:A47814, Thermo Scientific Corporation, Carlsbad, CA). The median Cq values (N-gene, Orf1ab, & S-gene (where applicable)) was calculated and used to determine how much the RNA needed to be diluted with water to reach a Cq value of 23. A post dilution RT-qPCR reaction was performed and used to calculate the final dilution of the more concentrated samples to the new target value of Cq 23.296. The number of freeze thaw cycles between RNA samples was kept the same.

#### Virus Mixing

RNA standardized in the prior section was used to make a volumetric mixing array (final volume 10uL) using a Mosquito X1 HV robotic liquid handler (SPT Labtech, UK). Pairwise mixes of 5:95, 10:90, 20:80, 60:40, and 50:50 were made for each virus lineage and in both directions. Equal mixes (20%) for each of the five test strains were made. 25% mixes and 33% mixes were made for a subset of possible combinations and controls of 100:0 were prepared. See **Extended Data Table 1** for complete array. Corrected estimates of the fraction of each virus lineage based were performed using the final measured Cq values for each pure virus lineage sample to control for issues encountered during the dilution step (repeat Cq measurements had a coefficient of variation of 0.007, **Extended Data Table 2**). Across all 95 mixtures, we observed a coefficient of variation of 0.016. Since initial virus concentrations are controlled for using measured Cq values, we expect remaining lineage specific bias (see **Extended Data Figure 3**) is likely due to experimental inconsistencies encountered during mixture creation.

### Deconvolution method performance comparison

A subset of the spike-in mixtures (1 of each type, for a total of 95 mixtures) was used to compare Freyja, cojac (using VOC definitions from the public cojac github repository; Lineage A and Epsilon definitions were created manually), the Kallisto-based method from Baaijens et al. 2021, and LCS. Kallisto was run using 10 cores (with no bootstrapping), and LCS was run using 16 cores, both on an Intel Xeon processor (2.2GHz). LCS was run for 48 hours, but failed to complete. Timing was performed using the “time” command, and included all steps after alignment, trimming, and sorting. Times correspond to total CPU time.

### Estimation of delay in detection frequency

Estimation of the lag time between epidemiological curves for wastewater and clinical surveillance of the Epsilon variant in San Diego was performed by identifying the shift with maximal cross-correlation. All time points leading up to the time of initial peak in detection frequency were included for both wastewater and clinical data.

### Phylogenetic analyses

Reconstruction of maximum likelihood trees was performed on all SARS-CoV-2 VOC genomes with 10x (10 reads or greater per site) genome coverage >95% and quality score >20 obtained from UCSD campus sampling, using IQtree^34^. This analysis included 150 (112 clinical, 38 wastewater) Epsilon, 49 (37 clinical, 12 wastewater) Alpha, and 160 (136 clinical, 24 wastewater) Delta lineage genomes from UCSD, in addition to 60 Epsilon, 20 Alpha, and 39 Delta randomly selected genomes from elsewhere in the United States. We used iVar ^20^ to identify consensus sequences for all San Diego samples. Bases were only included in the sequence if there was a consensus base at the site (>50% nucleotide frequency). We also masked known homoplasic sites prior to tree reconstruction^35^. Analysis of temporal comparison was performed on 608 samples (443 clinical, 165 wastewater, all lineages were included) with 10x genome coverage >95% and quality score >20 from UCSD. Sample collection SNP distances were calculated without considering ambiguous bases and gaps.

## Data Availability

Data availability and relevant links provided in the manuscript

https://searchcovid.info/dashboards/wastewater-surveillance/

## Code availability

Freyja is hosted publicly on github (https://github.com/andersen-lab/Freyja) and is available under a BSD-2-Clause License. Freyja is accessible as a package via bioconda (https://bioconda.github.io/recipes/freyja/README.html) in container form via dockerhub (https://hub.docker.com/r/andersenlabapps/freyja). COVID-19 VIral Epidemiology Workflow (C-VIEW) is available at https://github.com/ucsd-ccbb/C-VIEW as an open-source, end-to-end workflow for viral epidemiology focused on SARS-CoV-2 lineage assignment and phylogenetics.

## Data Availability

All raw wastewater sequencing data is available via the NCBI Sequence Read Archive under the BioProject ID PRJNA819090. Consensus sequences from clinical and wastewater surveillance are all available on GISAID. Spike-in sequencing data is available via google cloud (https://console.cloud.google.com/storage/browser/search-reference_data). The UCSD campus dashboard can be accessed at https://returntolearn.ucsd.edu/dashboard/. The county wastewater data from Point Loma are available through the public dashboard that can be accessed at https://searchcovid.info/dashboards/wastewater-surveillance/. The SEARCH genomic surveillance dashboard is available at https://searchcovid.info/dashboards/sequencing-statistics/.

## Acknowledgements

We thank Laralyn Asato and the Microbiology Lab at the SD Public Utilities Department for providing us with county wastewater samples. We thank UC San Diego’s Return to Learn (RTL) program for funding the campus-wide wastewater surveillance efforts. We also thank Jason Kayne, Rich Cota, Jesus Ortiz, and the Facilities management team (FM) at UCSD, Joseph Mayer from the Center for Aerosol Impacts on Chemistry of the Environment (CAICE) and Luke Arnold of the Campus Research Machine Shop (CRMS) for assistance with the installation and operation of the autosamplers; Robbie Jacobs, Shawn Knepple and their team at UCSD Logistics for assisting with our daily sampling efforts; Brett Pollak and the UCSD Information Technology Services team for assisting with the daily notifications; Office of Academic Affairs for contact tracing and targeted campus messaging assistance; Jana Severson, Patrick Hochstein, the UCSD HDH team, and the UCSD Environmental Health and Safety (EHS) personnel; Jack Gilbert and the Microbiome Sample Processing Core at UCSD for access to qPCR equipment. We also thank the CDC SPHERES consortium, SEARCH (San Diego Epidemiology and Research for COVID Health) Alliance, and members of the Andersen lab for discussion and help with logistics. We thank the healthcare workers, frontline workers, and patients who made the collection of this SARS-CoV-2 dataset possible and all those who made genomic data available for analysis via GISAID.

## Ethics declarations

The University of California San Diego Institutional Review Boards (IRB) provided human subject protection oversight of the of the data obtained by the EXCITE lab for the campus clinical samples (IRB approval #210699, #200477). All necessary patient/participant consent has been obtained and the appropriate institutional forms have been archived, and any sample identifiers included were de-identified. The wastewater component of this project was discussed with our Institutional Review Board, and was not deemed to be human subject research, as it did not record personally identifiable information.

## Author Contributions

Conceptualization: RK, KGA

Methodology: SK, JIL, NKM, PDH, AK, SS, KMF, AB, LCL, GWY, KGA, RK

Software: JIL, DM, NM, KMF, AB, BH, SS, KG, NLM, KSR, CMA, EH, AMM

Formal Analysis: SK, JIL

Investigation: SK, JIL, NM, SF, HMT, TV, CET, RT, NAB, TB, MC, WC, ESC, ERE, AH, GH, ALL, EL, TTN, TO, AP, RAS, PS, PBF, EWS, SA, PDH, CAM, LCL, GWY, CA, EK, MAS, SAP, JL, EP, MZ, ES, RFK, TG, RG, KGA, RK

Resources: CA, NKM, RMN, RS, EHS, AMS, SFK, DPD, CAH, AM, SS, BA, SS, NG, JDM, EM, IAM, AH, OB, AM, AB, KMSB, ETC, NLW, WL, MI, DB, LN, SW, MZ, RRS, RFM, TG, RG, DBA, DB, JCB, AB, ME, JG, SLG, MH, FKH, LI, HJ, TJ, VK, BK, LR, CAH, GCM, PM, RM, PO, DP, AP, AMS, BS, AS, BW, TW, SR, PKK ATY, DMC

Data curation: SK, JIL, PDH, GH, SF, HMT, CET, RT, TV, PDH, AB, NM, AMM, KMF

Writing – Original Draft: SK, JIL, KGA, RK

Writing – Review and editing: all authors

Visualization: SK, JIL

Supervision: RMN, NKM, RS, ALS, EHS, AMS, PDH, LCL, DP, GWY, KGA, RK

Project administration: RMN, NKM, RS, ALS, EHS, AMS, PDH, LCL, GWY, KGA, RK

Funding acquisition: RK, KGA

## Extended Data

**Extended Data Table 1:** Platemap of spike-in mixtures used for method validation.

|  | 1 | 2 | 3 | 4 | 5 | 6 |
| --- | --- | --- | --- | --- | --- | --- |
| **A** | 5% Delta: 95% A | 10% Delta: 90% A | 20% Delta: 80% A | 40% Delta: 60% A | 50% Delta: 50% A | 100% A |
| **B** | 5% Delta: 95% Beta | 10% Delta: 90% Beta | 20% Delta: 80% Beta | 40% Delta: 60% Beta | 50% Delta: 50% Beta | 100% Delta |
| **C** | 5% Delta: 95% Gamma | 10% Delta: 90% Gamma | 20% Delta: 80% Gamma | 40% Delta: 60% Gamma | 50% Delta: 50% Gamma | 100% Beta |
| **D** | 5% Delta: 95% Alpha | 10% Delta: 90% Alpha | 20% Delta: 80% Alpha | 40% Delta: 60% Alpha | 50% Delta: 50% Alpha | 100% Gamma |
| **E** | 5% Beta: 95% A | 10% Beta: 90% A | 20% Beta: 80% A | 40% Beta: 60% A | 50% Beta: 50% A | 100% Alpha |
| **F** | 5% Beta: 95% Delta | 10% Beta: 90% Delta | 20% Beta: 80% Delta | 40% Beta: 60% Delta | 50% Beta: 50% Delta | 20% A: 20% Delta: 20%<br>Beta: 20% Gamma: 20%<br>Alpha |
| **G** | 5% Beta: 95% Gamma | 10% Beta: 90% Gamma | 20% Beta: 80% Gamma | 40% Beta: 60% Gamma | 50% Beta: 50% Gamma | 25% Delta: 25% Beta<br>: 25% Gamma: 25%<br>Alpha |
| **H** | 5% Beta: 95% Alpha | 10% Beta: 90% Alpha | 20% Beta: 80% Alpha | 40% Beta: 60% Alpha | 50% Beta: 50% Alpha | 25% Delta: 25% Beta:<br>25% Gamma: 25% A |
| **I** | 5% Gamma: 95% A | 10% Gamma: 90% A | 20% Gamma: 80% A | 40% Gamma: 60% A | 50% Gamma: 50% A | 25% Delta: 25% Beta:<br>25% A: 25% Alpha |
| **J** | 5% Gamma: 95% Delta | 10% Gamma: 90% Delta | 20% Gamma: 80% Delta | 40% Gamma: 60% Delta | 50% Gamma: 50% Delta | 25% Delta: 25% A: 25%<br>Gamma: 25% Alpha |
| **K** | 5% Gamma: 95% Beta | 10% Gamma: 90% Beta | 20% Gamma: 80% Beta | 40% Gamma: 60% Beta | 50% Gamma: 50% Beta | 25% A: 25% Beta: 25%<br>Gamma: 25% Alpha |
| **L** | 5% Gamma: 95% Alpha | 10% Gamma: 90% Alpha | 20% Gamma: 80% Alpha | 40% Gamma: 60% Alpha | 50% Gamma: 50% Alpha | 33% Delta: 33% Beta:<br>33% Gamma |
| **M** | 5% Alpha: 95% A | 10% Alpha: 90% A | 20% Alpha: 80% A | 40% Alpha: 60% A | 50% Alpha: 50% A | 33% Delta: 33% Beta:<br>33% Alpha |
| **N** | 5% Alpha: 95% Delta | 10% Alpha: 90% Delta | 20% Alpha: 80% Delta | 40% Alpha: 60% Delta | 50% Alpha: 50% Delta | 33% Delta: 33% Alpha:<br>33% Gamma |
| **O** | 5% Alpha: 95% Beta | 10% Alpha: 90% Beta | 20% Alpha: 80% Beta | 40% Alpha: 60% Beta | 50% Alpha: 50% Beta | 33% Alpha: 33% Beta:<br>33% Gamma |
| **P** | 5% Alpha: 95% Gamma | 10% Alpha: 90% Gamma | 20% Alpha: 80% Gamma | 40% Alpha: 60% Gamma | 50% Alpha: 50% Gamma | Neg |

**Extended Data Table 2:** Consistency of Lineage A Cq values across repeated measurements.

| **Replicate Number** | **Cq N Gene** | **Cq Orflab** | **Cq S Gene** | **Cq RNaseP** | **Average** |
| --- | --- | --- | --- | --- | --- |
| 1 | 31.228 | 30.807 | 30.045 | 29.581 | 30.693 |
| 2 | 30.783 | 29.77 | 29.546 | 29.49 | 30.033 |
| 3 | 31.201 | 30.622 | 29.733 | 29.745 | 30.519 |
| 4 | 30.621 | 30.953 | 29.578 | 28.925 | 30.384 |
| 5 | 31.188 | 30.073 | 29.366 | 28.745 | 30.209 |
| 6 | 30.604 | 29.788 | 29.829 | 28.797 | 30.074 |
| 7 | 30.308 | 30.335 | 29.573 | 29.149 | 30.072 |
| 8 | 30.738 | 30.36 | 29.711 | 28.79 | 30.269 |
| 9 | 31.144 | 29.97 | 30.045 | 28.79 | 30.386 |
| 10 | 31.122 | 30.822 | 29.566 | 29.671 | 30.503 |
| 11 | 31.825 | 29.763 | 29.833 | 29.134 | 30.474 |
| 12 | 31.434 | 30.18 | 29.773 | 29.133 | 30.462 |
| 13 | 31.209 | 29.793 | 29.402 | 29.559 | 30.135 |
| 14 | 30.641 | 30.181 | 29.816 | 29.833 | 30.213 |
| 15 | 30.744 | 29.371 | 29.695 | 29.257 | 29.937 |
| 16 | 30.396 | 29.728 | 29.441 | 28.428 | 29.855 |
| 17 | 30.957 | 29.449 | 29.913 | 28.242 | 30.107 |
| 18 | 30.791 | 30.113 | 29.601 | 29.277 | 30.169 |
| 19 | 31.561 | 29.839 | 29.943 | 29.06 | 30.448 |
| 20 | 31.434 | 29.711 | 29.568 | 28.864 | 30.238 |

**Extended Data Table 3:** Omicron surveillance at Point Loma Wastewater Treatment Plant.

| Collection Date | Avg. Estimated Omicron Abundance (%) | qPCR Detection |  |  |
| --- | --- | --- | --- | --- |
|  |  | DelHV69/70 | N501Y | P681R |
| 10/04/21 | 0 |  |  | x |
| 10/06/21 | 0 |  |  | x |
| 10/10/21 | 0 |  |  | x |
| 10/11/21 | 0 |  |  | x |
| 10/13/21 | 0 |  |  | x |
| 10/17/21 | 0 |  |  | x |
| 10/18/21 | 0 |  |  | x |
| 10/20/21 | 0 |  |  | x |
| 11/12/21 | 0 |  |  | x |
| 11/22/21 | 0 |  |  | x |
| 11/27/21 | 1.726 | x | x | x |
| 11/28/21 | 1.967 | x | x | x |
| 12/1/21 | 2.439 | x | x | x |
| 12/5/21 | 17.11 | x | x | x |
| 12/6/21 | 19.764 | x | x | x |
| 12/12/21 | 50.65 | x | x | x |
| 12/16/21 | 67.14 | x | x | x |
| 12/20/21 | 79.135 | x | x | x |
| 12/21/21 | 80.567 | x | x | x |

**Extended Data Figure 1:**
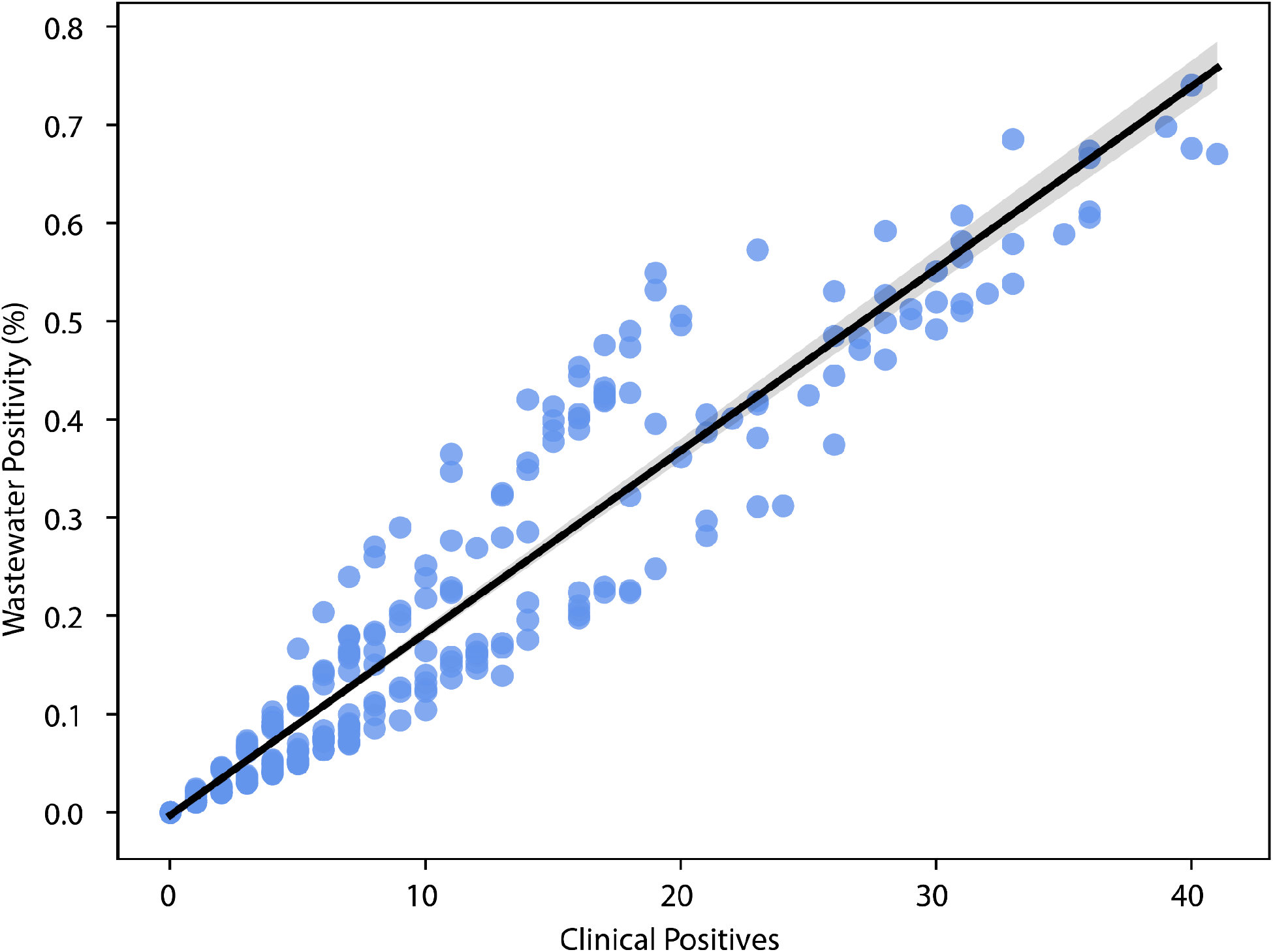
Relationship of daily UCSD campus wastewater sampler positivity and campus clinical positives. Black line indicates the linear fit to the data, with bootstrap 95% confidence interval shown in gray.

**Extended Data Figure 2:**
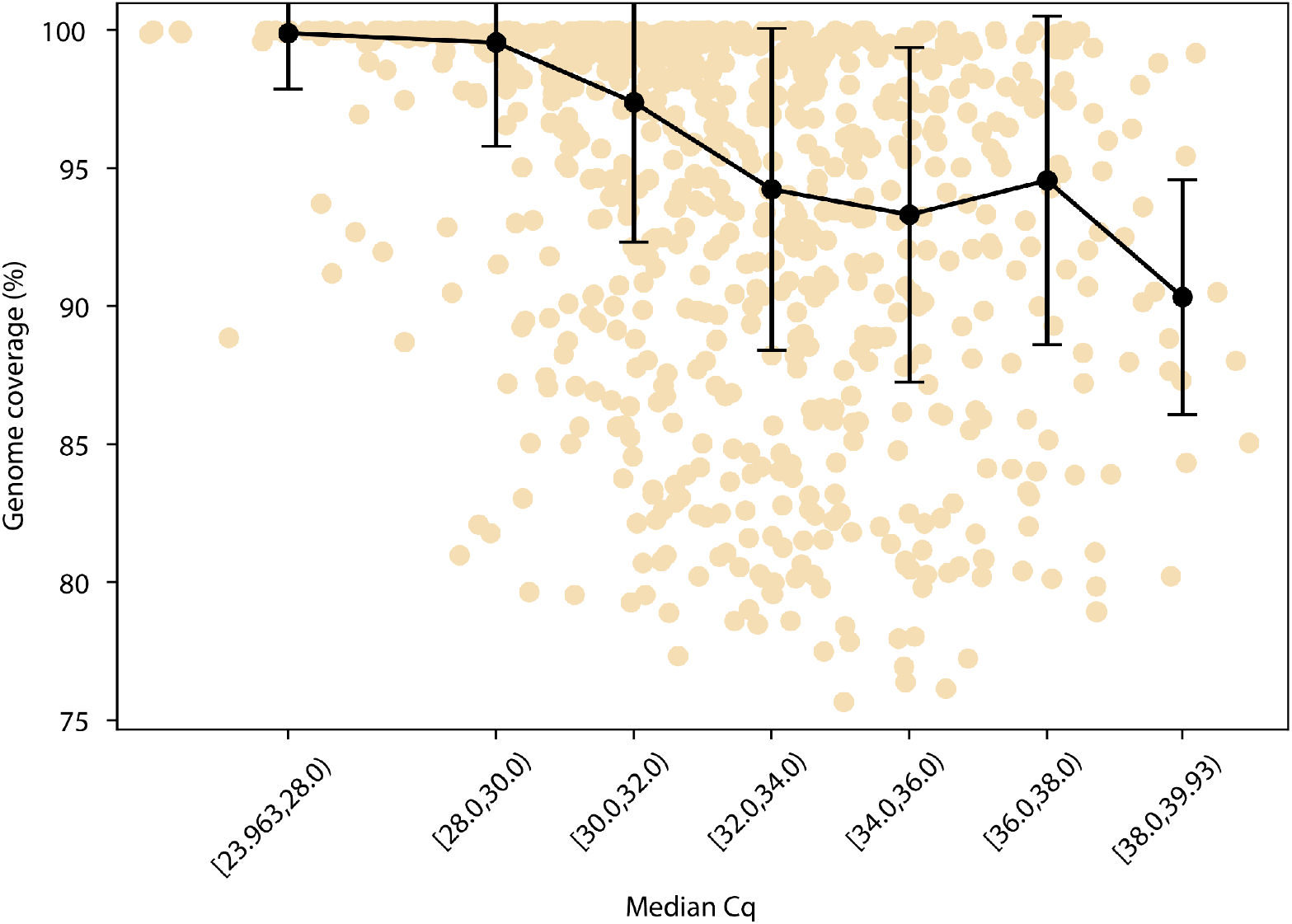
Relationship between genome coverage and cycle quantification values. 10x genome coverage (fraction of sites with 10 reads or greater) remains high, even for Cq values of nearly 38. Points indicate median value in each bin, while error bars indicate the median absolute deviation.

**Extended Data Figure 3:**
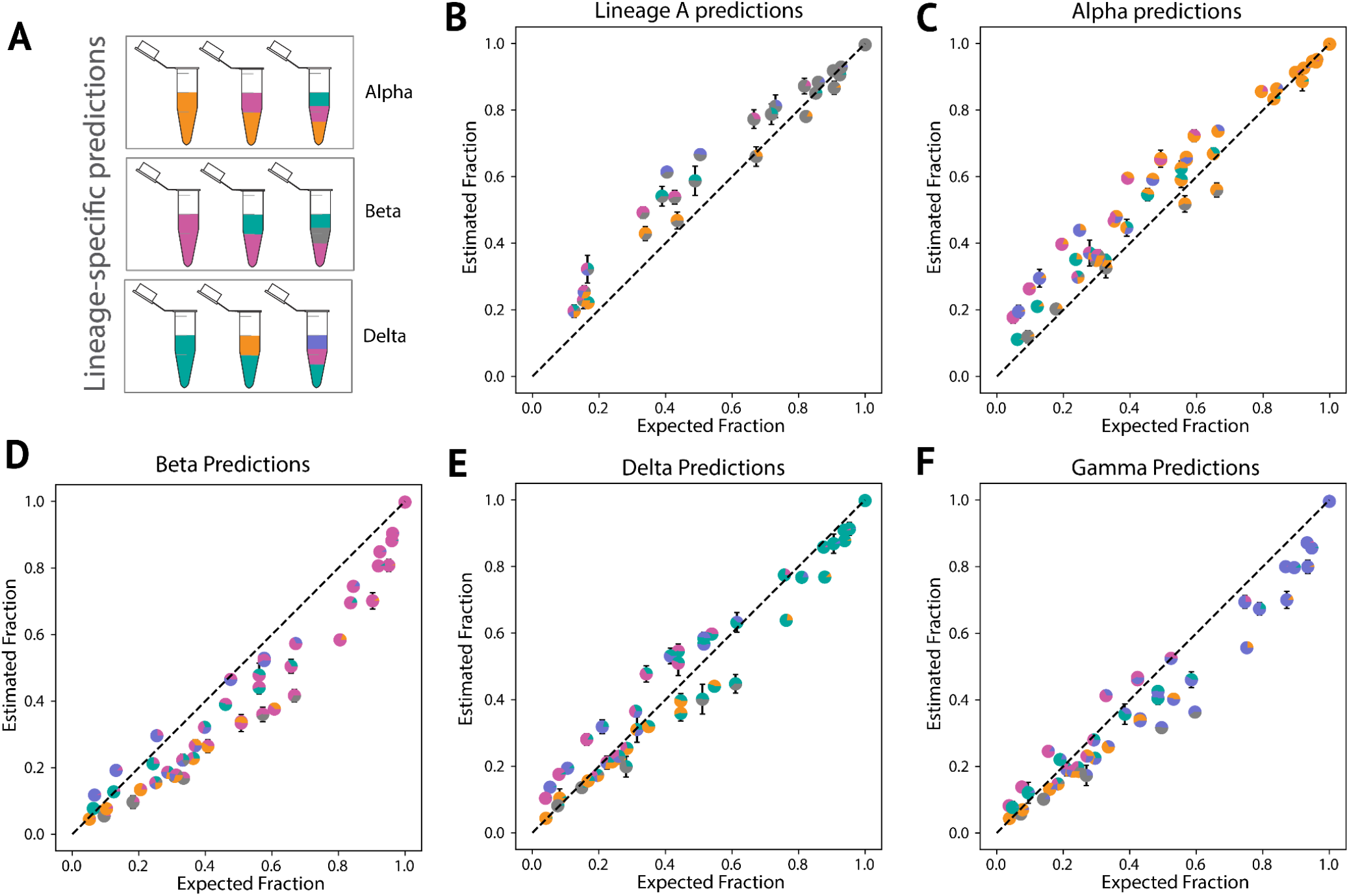
Lineage-specific prediction of variant abundance in spike-in validation samples. A. Schematic of “spike-in” sample design. B-F. Lineage specific prediction. Proportions of each lineage in the sample are shown as a pie chart marker (Grey = Lineage A, Orange = Alpha, Pink = Beta, Turquoise = Delta, and Purple = Gamma) with error bars indicating the standard deviation from the mean, across four replicates.

**Extended Data Figure 4:**
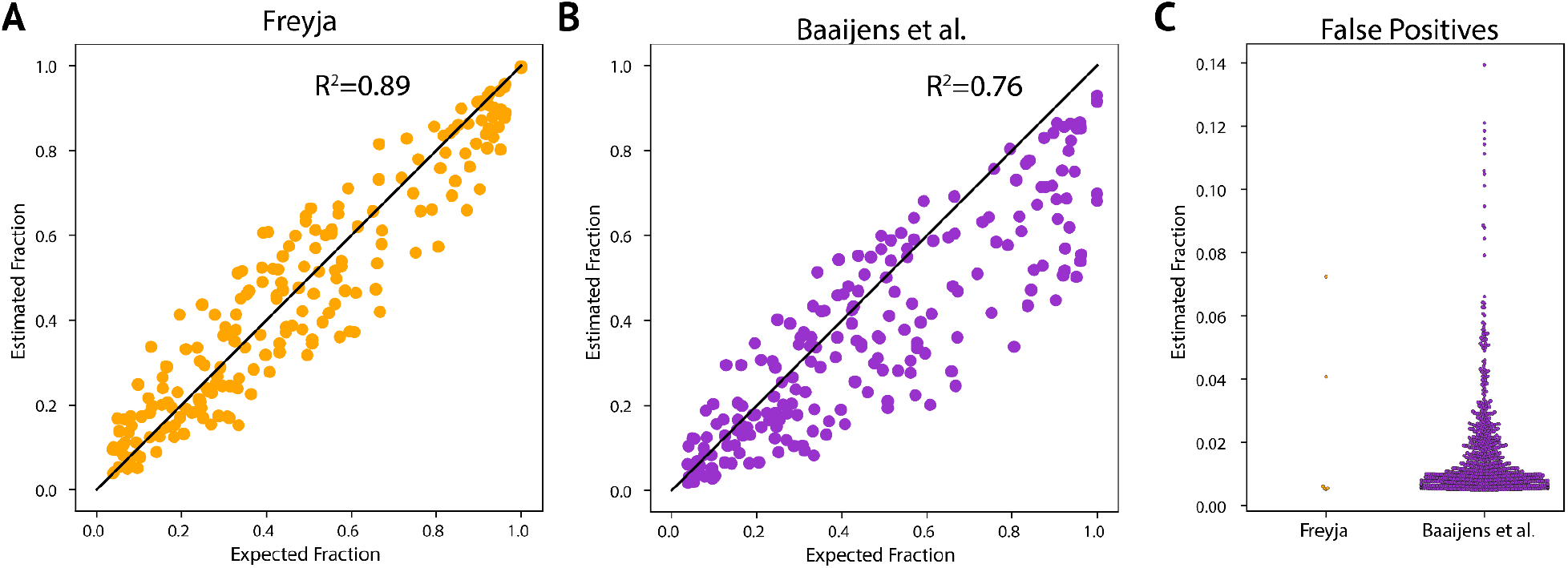
Freyja more accurately estimates virus abundance, with fewer false positives. A-B. Estimated vs expected fraction of each lineage in the mixture. The Kallisto-based approach from Baaijens et. al shows a wider range of estimates for each known mix fraction, and generally underestimates the fraction. C. False positives with abundance greater than 0.5%.

**Extended Data Figure 5:**
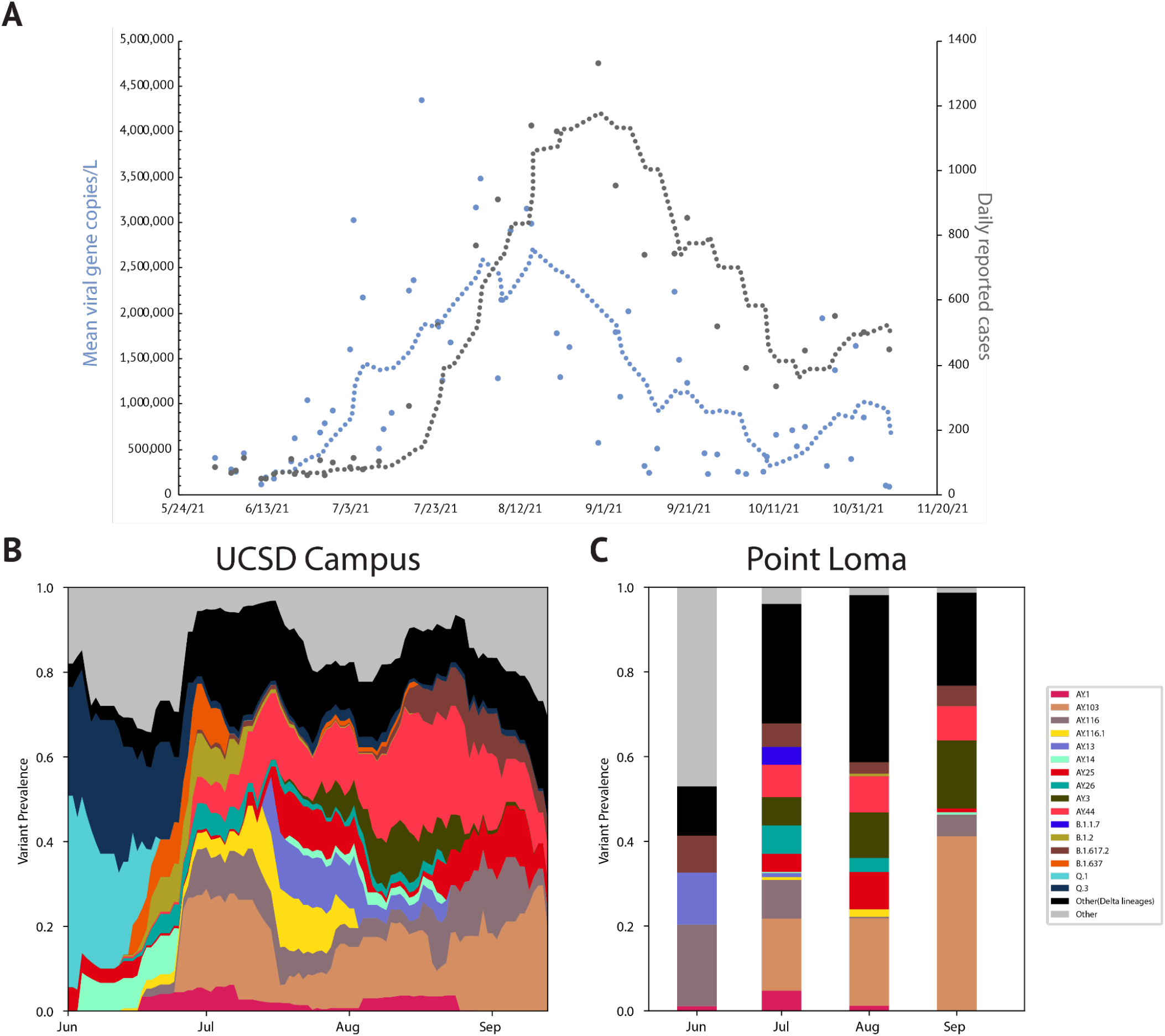
The rise of the Delta variant during Summer 2021. A. Mean SARS-CoV-2 viral gene copies/L of raw sewage (blue) collected from the Point Loma Wastewater Treatment Plant and caseload (gray) reported by the county during the same period. SARS-CoV-2 concentrations were normalized by PMMoV (pepper mild mottle virus) concentration to adjust for load changes. B. Lineage distribution in UCSD campus wastewater. C. Monthly lineage averages for wastewater collected at Point Loma Wastewater Treatment Plant during the Delta surge (N= 5, 20, 25, 7)

**Extended Data Figure 6:**
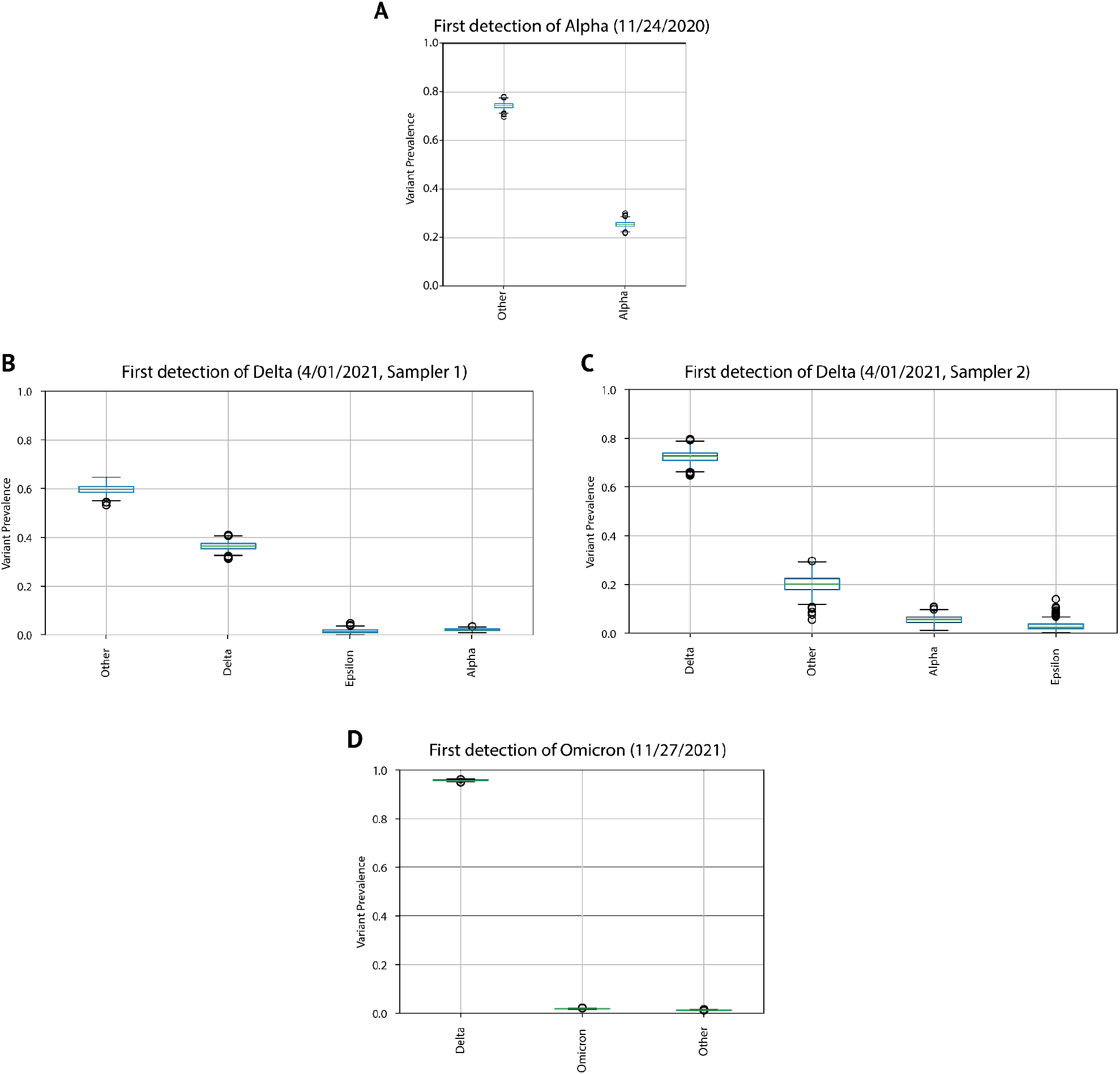
Quantification of deconvolution uncertainty in first detection of VOCs. A-D. Bootstrap distributions of Freyja abundance estimates obtained by resampling read data from each sample corresponding to the first detection of that VOC in San Diego. Two samplers were found to contain Delta on the same day. First detections were also confirmed using a VOC qPCR panel, as shown in Figure 2 and Extended Data Table 3. 95% Confidence intervals for variant prevalence for each first detection event: A. Alpha: (0.232, 0.278), B. Delta: (0.336, 0.397), C. Delta: (0.676, 0.772), D. Omicron: (0.017, 0.021).

**Extended Data Figure 7:**
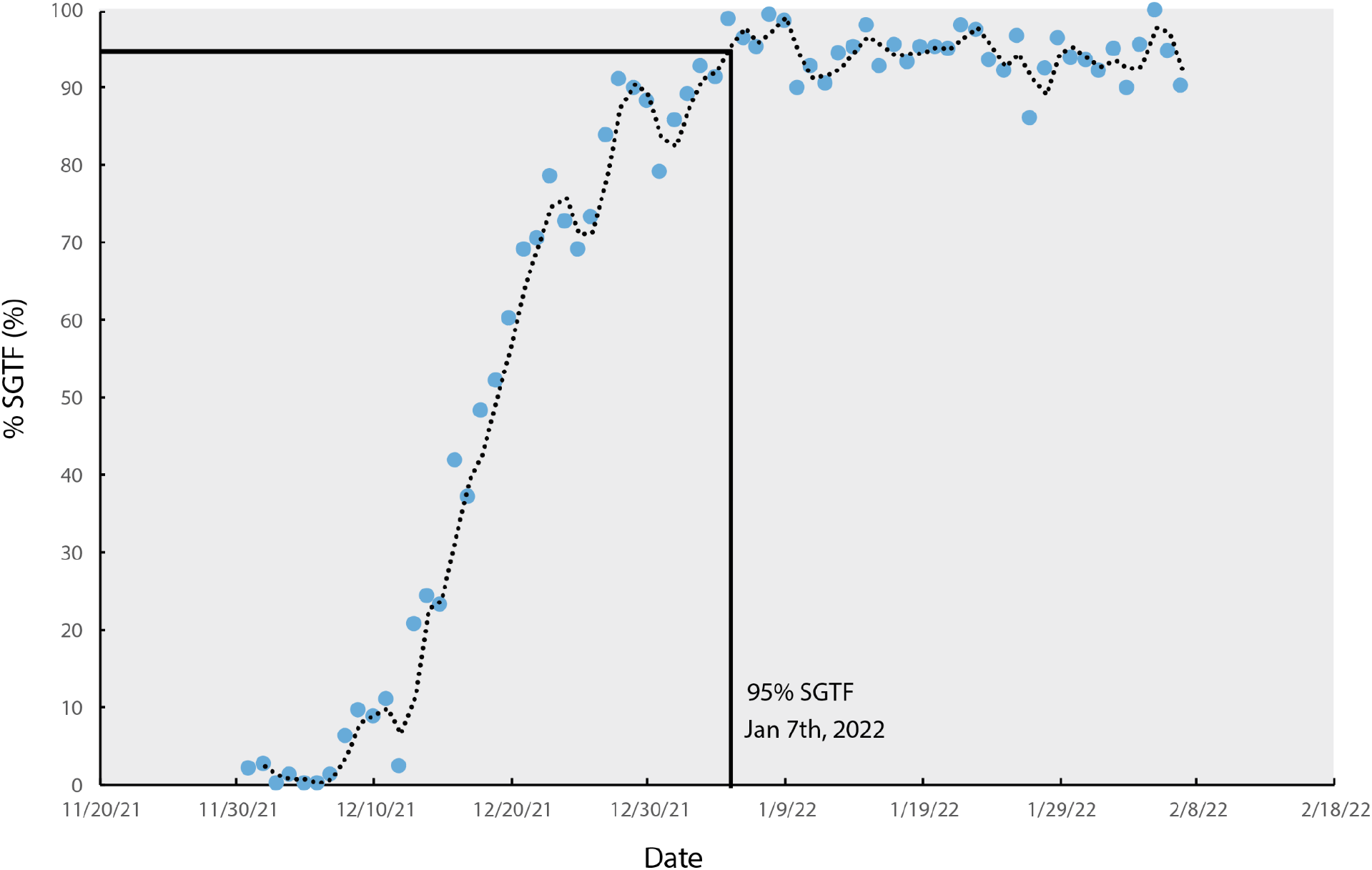
Estimated proportion of Omicron sequences in clinical data. Omicron estimates tracked via S-gene target failure, SGTF (characteristic of Omicron lineage BA.1 and its descendants) qPCR assays for clinical samples in San Diego between November 27th, 2021-February 7th, 2022. First detection of Omicron through clinical genomic sequencing in San Diego was December 8th. Dotted line shows a rolling average with a window size of seven days.

